# Modifiable and Context-Specific Factors Associated with Dementia in the Democratic Republic of the Congo: A Community-Derived Cross-Sectional Study

**DOI:** 10.64898/2026.11.16.26368211

**Authors:** Jean Ikanga, Saranya Sundaram Patel, Peng Zhang, Jingjing Yang, Caterina Obenauf, Guy Gikelekele, Emmanuel Epenge, Japhet Magolu Potshi, Topina Tomadia, Joseph Phuati Tsangu, Samuel Mampunza, Lelo Mananga, Justine Bukabau, Thomas Karikari, Alden L. Gross, Alvaro Alonso

**Affiliations:** Emory University School of Medicine, Department of Rehabilitation Medicine, Atlanta, GA 30322, USA; University of Kinshasa and Catholic University of Congo, School of Medicine, Department of Psychiatry, B.P. 7463 Kinshasa I, Democratic Republic of the Congo; One Rehab, Dallas, TX, USA; Emory University School of Medicine, Department of Human Genetics, Atlanta, GA 30322, USA; University of Tennessee, Department of Psychology and Neuroscience, Knoxville, TN, USA; University of Kinshasa, Department of Neurology, B.P. 7463 Kinshasa I, Democratic Republic of the Congo; Protestant University of Congo, School of Medicine, Kinshasa, Democratic Republic of the Congo; Memory Clinic of Kinshasa, B.P. 7463 Kinshasa I, Democratic Republic of the Congo; University of Kinshasa, Department of Anesthesiology and Reanimation, B.P. 7463 Kinshasa, Democratic Republic of the Congo; University of Kinshasa, Department of Internal Medicine, B.P. 7463 Kinshasa I, Democratic Republic of the Congo; University of Pittsburgh School of Medicine, Department of Psychiatry, Pittsburgh, PA, USA; Johns Hopkins Bloomberg School of Public Health, Department of Epidemiology, Baltimore, MD, USA; Department of Preventive Medicine and Public Health, School of Medicine, University of Navarra, Pamplona, Spain

**Author notes:** Corresponding author: Jean N. Ikanga, Ph.D. | Department of Rehabilitation Medicine, Emory University | 1441 Clifton Rd NE, Atlanta, GA 30322, USA |.

**Keywords:** Alzheimer’s disease, Dementia, Democratic Republic of the Congo, Social Determinants of Health, Logistic Models, Cognitive Aging

## Abstract

**Background:** Evidence linking social, behavioral, and environmental factors to dementia in Central Africa remains limited.

**Objective:** To examine associations between 16 factors and dementia status in the Democratic Republic of the Congo (DRC), and quantify incremental model fit beyond age and sex.

**Methods:** This cross-sectional analysis included 721 participants (232 cognitively unimpaired; 489 with dementia, including 247 with suspected Alzheimer’s disease). Logistic regression jointly modeled 16 factors, age, sex, and missing-category indicators. Continuous covariates with missing values were mean-imputed and standardized. Model fit was assessed using Nagelkerke pseudo-R², likelihood-ratio testing, and 1,000 bootstrap replicates.

**Results:** Nagelkerke pseudo-R² increased from 0.106 with age and sex to 0.368 after adding the factors (ΔR²=0.262; bootstrap 95% CI 0.241–0.371; χ²(27)=163.62, p<0.001). Poverty (adjusted odds ratio [aOR] 2.34, 95% CI 1.52–3.60), low resilience (aOR 1.84, 95% CI 1.18–2.86), and depression (aOR 1.53, 95% CI 1.02–2.31) were associated with higher dementia odds. Higher education (aOR 0.51, 95% CI 0.39–0.66, per standard-deviation increase) and obesity (aOR 0.31, 95% CI 0.10–0.90) showed inverse associations; obesity data were 79.9% missing. Alcohol consumption and PM₂.₅ exposure were not significantly associated with dementia.

**Conclusions:** Poverty, lower resilience, depression, and education were associated with dementia status in this nonrepresentative cohort. Incremental model fit does not quantify preventable dementia burden, and causality cannot be inferred from these cross-sectional data.

## INTRODUCTION

Dementia is an increasing public health concern as populations age, with particularly large projected increases in low- and middle-income countries.^1^ The 2024 Lancet Commission emphasized that many established exposures are potentially modifiable over the life course.^2^ However, the prevalence, timing, and interrelationship of dementia-associated exposures vary substantially across populations. Findings derived mainly from high-income countries may not capture the circumstances of older adults in sub-Saharan Africa.^3,4^

Studies from African settings underscore the potential relevance of educational disadvantage, depression, and cardiometabolic health, while highlighting variation across countries.^5,6^ Structural circumstances may also shape late-life cognition through interrelated pathways involving childhood educational opportunity, cumulative material deprivation, access to care, nutrition, environmental hazards, and exposure to chronic stress. The lived experience of conflict and displacement may have consequences for health and functioning that persist across the life course.^7,8^

The Democratic Republic of the Congo (DRC) is a particularly informative setting for examining factors associated with dementia. In addition to established vascular and lifestyle exposures, economic insecurity, educational inequality, trauma, and experiences related to armed conflict may influence older adults’ cognitive and functional health.^9,10^ The EVCD-RDC cohort combines culturally adapted cognitive assessment with clinical adjudication and measures of social and environmental exposures.^11,12^ Nevertheless, the cross-sectional nature of baseline data and selection through community and clinical settings call for particular caution when interpreting associations.

Population-attributable fractions require assumptions about causal effects, representative exposure distributions, and interpretable risk estimates that are not established by the present cross-sectional, dementia-enriched sample.^3,4^ Accordingly, this study focuses on mutually adjusted associations between the 16 factors and dementia status, and the incremental improvement in logistic model fit after adding these factors to age and sex.

## METHODS

### Study design and participants

This cross-sectional analysis used baseline data from the Étude du Vieillissement Cognitif et de Démence en République Démocratique du Congo (EVCD-RDC), a community-derived cohort investigating cognitive aging and dementia in the DRC.^11^ Recruitment through community organizations and clinical sites in Kinshasa was not based on probability sampling. Accordingly, the analytic group should not be considered representative of the older DRC population.

For the present analysis, participants with a documented age younger than 60 years were excluded. Starting with 1,044 unique baseline records, exclusions were applied sequentially: 268 with mild cognitive impairment (MCI), 41 without a confirmed cognitively unimpaired (CU) or dementia classification, one with known age younger than 60 years, and 13 dementia cases with missing information on depression, social isolation, civilian war exposure, or traumatic experience. The latter exclusion was made because outcome-specific missing categories could produce separation and unstable logistic estimates. The final sample included 721 participants (232 CU; 489 dementia, comprising 247 suspected Alzheimer’s disease and 242 other dementias). Two participants without a valid age had their ages imputed by the cohort mean and were included in the analyses.

As described in the EVCD-RDC cohort methods,^11^ neurocognitive and functional evaluations were conducted by trained personnel and diagnoses were adjudicated by a multidisciplinary clinical panel. CU required preserved cognition and everyday functioning; dementia classification incorporated multidomain cognitive impairment with functional decline. The IDEA instrumental activities of daily living assessment was part of diagnostic evaluation.^13,14^ Etiologic categories were assigned from the available cognitive, medical, and neurological information, and should be regarded as clinical classifications rather than biomarker-confirmed Alzheimer’s disease.

### Exposures and measures

The 16 potentially modifiable or context-specific factors were education (continuous years), smoking, alcohol consumption, diabetes, sleep disorder, hypertension, obesity, depression, social isolation, poverty, civilian war experience, traumatic events, higher particulate matter exposure (PM₂.₅), hearing loss, low resilience, and low literacy (Table 1). These measures were obtained from the National Alzheimer’s Coordinating Center Uniform Data Set, culturally adapted instruments, study questionnaires, and brief environmental monitoring.^11,15^ The exposures include both established potentially modifiable risk factors and contextual indicators that could be modifiable in earlier life. Poverty was measured using an adapted multidimensional deprivation index that includes education and health-related dimensions and may overlap with other modeled exposures. Social isolation was operationalized as living alone, whereas civilian war experience and other traumatic events were assessed separately. Resilience and literacy were represented by dichotomized questionnaire scores. Higher PM₂.₅ exposure was classified using a cutoff of >15 µg/m³ applied to approximately one-hour indoor ATMOtube PRO measurements, which were not obtained for all participants. Obesity and hypertension measurements were introduced after the initial study phase, resulting in 79.9% missing obesity data and 39.4% missing hypertension data. Operational definitions and missingness are detailed in Table 1.

**Table 1.**
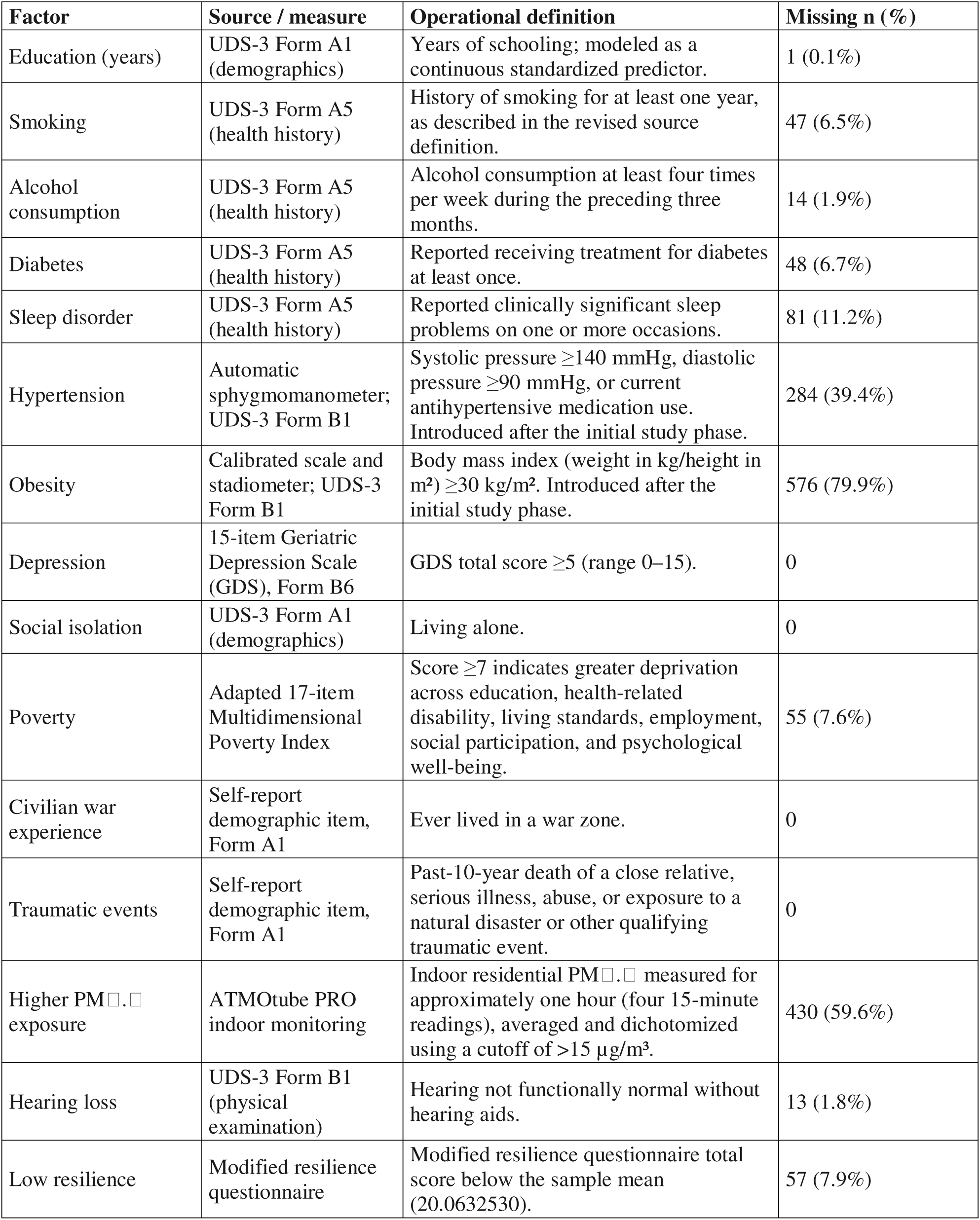
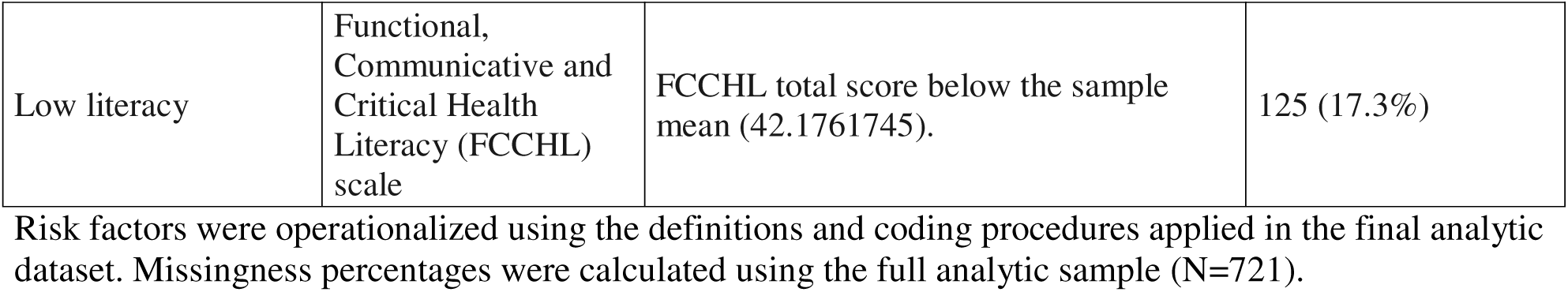
Definition and measurement of dementia-related factors and final-sample missingness.

| Factor | Source / measure | Operational definition | Missing n (%) |
|---|---|---|---|
| Education (years) | UDS-3 Form A1 (demographics) | Years of schooling; modeled as a continuous standardized predictor. | 1 (0.1%) |
| Smoking | UDS-3 Form A5 (health history) | History of smoking for at least one year, as described in the revised source definition. | 47 (6.5%) |
| Alcohol consumption | UDS-3 Form A5 (health history) | Alcohol consumption at least four times per week during the preceding three months. | 14 (1.9%) |
| Diabetes | UDS-3 Form A5 (health history) | Reported receiving treatment for diabetes at least once. | 48 (6.7%) |
| Sleep disorder | UDS-3 Form A5 (health history) | Reported clinically significant sleep problems on one or more occasions. | 81 (11.2%) |
| Hypertension | Automatic sphygmomanometer; UDS-3 Form B1 | Systolic pressure ≥140 mmHg, diastolic pressure ≥90 mmHg, or current antihypertensive medication use. Introduced after the initial study phase. | 284 (39.4%) |
| Obesity | Calibrated scale and stadiometer; UDS-3 Form B1 | Body mass index (weight in kg/height in m²) ≥30 kg/m². Introduced after the initial study phase. | 576 (79.9%) |
| Depression | 15-item Geriatric Depression Scale (GDS), Form B6 | GDS total score ≥5 (range 0–15). | 0 |
| Social isolation | UDS-3 Form A1 (demographics) | Living alone. | 0 |
| Poverty | Adapted 17-item Multidimensional Poverty Index | Score ≥7 indicates greater deprivation across education, health-related disability, living standards, employment, social participation, and psychological well-being. | 55 (7.6%) |
| Civilian war experience | Self-report demographic item, Form A1 | Ever lived in a war zone. | 0 |
| Traumatic events | Self-report demographic item, Form A1 | Past-10-year death of a close relative, serious illness, abuse, or exposure to a natural disaster or other qualifying traumatic event. | 0 |
| Higher PM□.□ exposure | ATMOtube PRO indoor monitoring | Indoor residential PM□.□ measured for approximately one hour (four 15-minute readings), averaged and dichotomized using a cutoff of >15 µg/m³. | 430 (59.6%) |
| Hearing loss | UDS-3 Form B1 (physical examination) | Hearing not functionally normal without hearing aids. | 13 (1.8%) |
| Low resilience | Modified resilience questionnaire | Modified resilience questionnaire total score below the sample mean (20.0632530). | 57 (7.9%) |
| Low literacy | Functional, Communicative and Critical Health Literacy (FCCHL) scale | FCCHL total score below the sample mean (42.1761745). | 125 (17.3%) |

Risk factors were operationalized using the definitions and coding procedures applied in the final analytic dataset. Missingness percentages were calculated using the full analytic sample (N=721).

### Statistical analysis

Age was calculated from the interval between the visit date and birth date (divided by 365), with a recorded age variable used when necessary. For the final sample, two missing ages and one missing education value were replaced by cohort means before standardization. Age and education were entered as standardized continuous variables. Binary factor coding used “No” as the reference, with separate “Yes” and, where values were unavailable, “Missing” indicators. Thus, absent exposure data were not recoded as a negative exposure. Sex was observed for all 721 participants.

The dichotomous outcome was dementia (suspected Alzheimer’s disease or other dementia) versus CU, which was the outcome in the considered logistic regression models. We fitted a baseline logistic regression model (M0) containing covariates of standardized age and sex, and a full model (M1) containing covariates of age, sex, and 16 modifiable factors considered in this study. Missing values were considered as an additional category in the categorical factors, which were considered as dummy variables in the logistic regression model with an additional missing indicator variable for each of the factors that have missing values. We also fitted an intercept-only logistic regression model for calculating the Nagelkerke pseudo-R²,^16^ which measures how model likelihood is improved over the likelihood of the intercept-only null model on a scale of 0 to 1. Let *L_0_* denote the likelihood of the intercept only null model; *L_M_* denote the likelihood of the considered model; *n* denote the sample size. The Nagelkerke pseudo-R² is calculated by 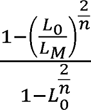.

Additionally, we fitted a sensitivity model (M2) by omitting obesity and PM₂.₅ factors with high missing rate from the full model. All models used the same 721 participants. Nagelkerke pseudo-R² values were calculated by comparing the loglikelihood of each fitted model to the intercept-only null model of the same cohort data. Incremental pseudo-R² was defined as ΔR² = R²(M1) − R²(M0), representing the amount of pseudo-R² attributed by adding these 16 modifiable factors. This ΔR² is an in-sample measure of the goodness of model fit, not the proportion of dementia cases caused by or preventable through the exposures. Likelihood-ratio test was conducted to compare M1 vs. M0.

Confidence intervals of Nagelkerke pseudo-R² and ΔR² were estimated by the nonparametric bootstrap of 1,000 replicates. In each bootstrap replicate, 721 participants were sampled from observed cohort data with replacement, and the resulting samples were used to fit the logistic regression models of intercept-only, baseline, full, and sensitivity models. Nagelkerke pseudo-R² value was calculated for each fitted model from each bootstrap replicate, and ΔR² was calculated for each compared model pairs (full vs. baseline, full vs. sensitivity). The 95% confidence intervals of the Nagelkerke pseudo-R² and ΔR² were derived from the 2.5th and 97.5th percentiles of their corresponding values obtained from 1,000 bootstrap replicates. Adjusted odds ratios (aORs) and the corresponding 95% confidence intervals for these 16 modifiable factors were estimated from the joint full model. Reported statistical significance is nominal and not corrected for multiple comparisons.

Analyses were conducted in R version 4.5.3 (R Foundation for Statistical Computing, Vienna, Austria).

### Ethics

The EVCD-RDC study protocol was approved by the Research Ethics Committee of the School of Public Health at the University of Kinshasa (original protocol ESP/CE/054c/2024; renewed July 18, 2026, through July 17, 2027) and the Emory University Institutional Review Board (IRB ID MOD006-IRB00113463). Written informed consent was obtained before participation, and study procedures were conducted in accordance with applicable ethical standards and the Declaration of Helsinki.

## RESULTS

### Participant characteristics

The final analytic sample included 721 participants, of whom 232 (32.2%) were cognitively unimpaired (CU) and 489 (67.8%) had dementia (247 with suspected Alzheimer’s disease and 242 with other dementia; Figure 1). The proportion of dementia cases reflects cohort selection and should not be interpreted as population prevalence. Median age was 72.5 years (IQR, 8.6) overall, 70.2 years (IQR, 7.1) among CU participants, and 73.7 years (IQR, 7.9) among participants with dementia. Women accounted for 462 participants (64.1%). Median years of education were 10.0 (IQR, 6.0) overall, 12.0 (IQR, 5.0) among CU participants, and 8.0 (IQR, 7.0) among participants with dementia (Table 2).

**Figure 1.**
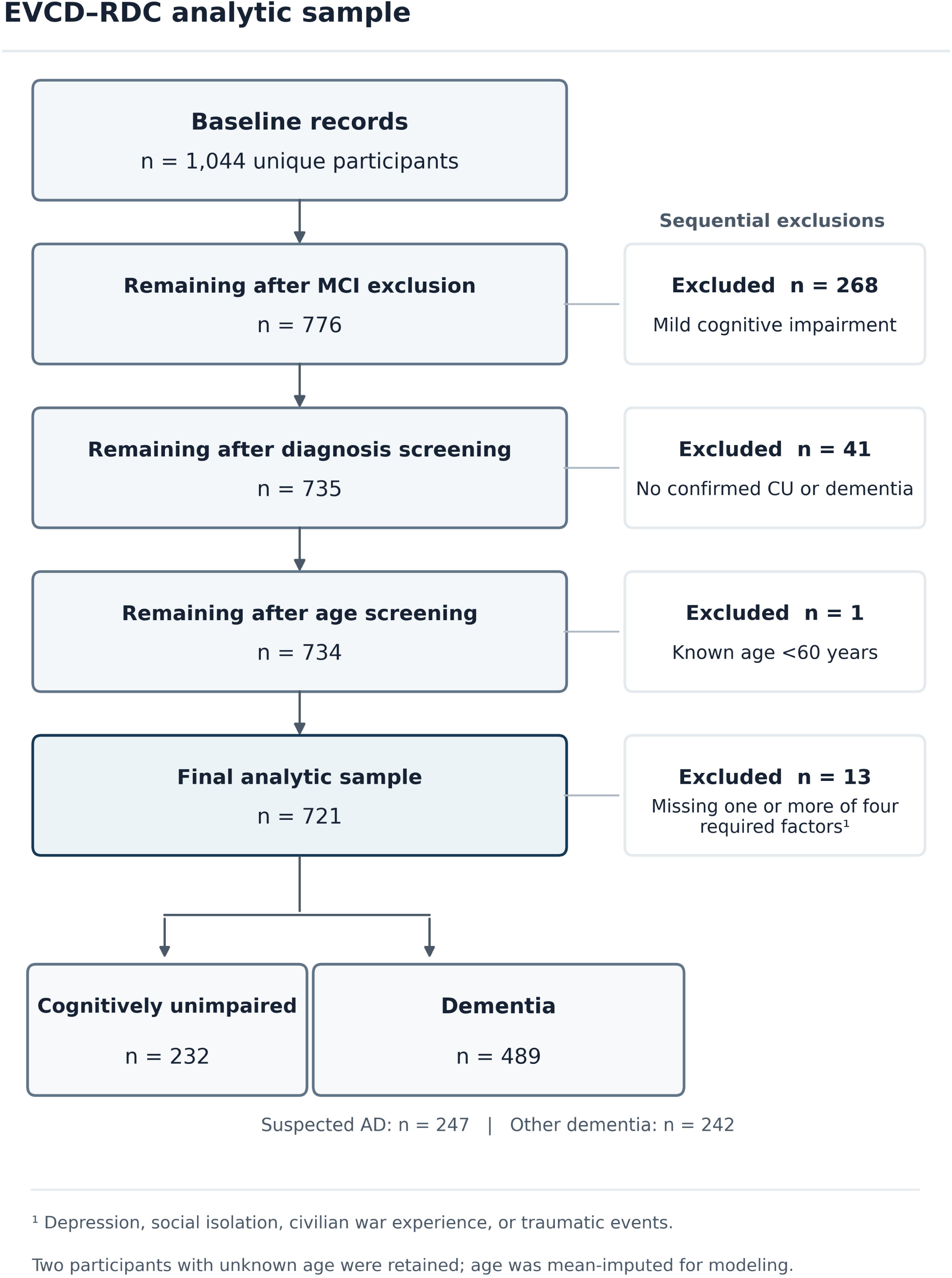
Selection of the 721-person EVCD-RDC analytic sample from 1,044 unique baseline records. The analytic eligibility rule excluded known ages below 60 years. Two participants with unknown age were retained with mean imputation; the cohort’s original published age eligibility should not be conflated with the analysis-specific cutoff. CU, cognitively unimpaired; MCI, mild cognitive impairment; AD, suspected Alzheimer’s disease.

**Table 2.** Characteristics of the analytic sample by cognitive status.

| **Characteristic** | **Overall (N=721)** | **CU (n=232)** | **Dementia (n=489)** |
|---|---|---|---|
| Demographics |  |  |  |
| Age, median (IQR) | 72.5 (8.6) | 70.2 (7.1) | 73.7 (7.9) |
| Female | 462 (64.1%) | 139 (59.9%) | 323 (66.1%) |
| Cognitive and Functional Assessments |  |  |  |
| MOCA | 20.0 (7.0) | 25.0 (4.0) | 18.0 (6.0) |
| CDR-SB | 2.8 (4.5) | 1.5 (2.0) | 3.5 (5.5) |
| IDEA-IADL | 27.0 (10.0) | 30.0 (7.0) | 25.0 (11.0) |
| Modifiable Risk Factors |  |  |  |
| Education, median (IQR) years | 10.0 (6.0) | 12.0 (5.0) | 8.0 (7.0) |
| Smoking | 47 (6.5%) | 21 (9.1%) | 26 (5.3%) |
| Alcohol Consumption | 15 (2.1%) | 7 (3.0%) | 8 (1.6%) |
| Diabetes | 59 (8.2%) | 15 (6.5%) | 44 (9.0%) |
| Sleep Disorder | 241 (33.4%) | 60 (25.9%) | 181 (37.0%) |
| Hypertension | 406 (56.3%) | 130 (56.0%) | 276 (56.4%) |
| Obesity | 32 (4.4%) | 10 (4.3%) | 22 (4.5%) |
| Depression | 312 (43.3%) | 75 (32.3%) | 237 (48.5%) |
| Social Isolation | 44 (6.1%) | 23 (9.9%) | 21 (4.3%) |
| Poverty | 285 (39.5%) | 51 (22.0%) | 234 (47.9%) |
| War Experience | 300 (41.6%) | 90 (38.8%) | 210 (42.9%) |
| Traumatic Events | 423 (58.7%) | 126 (54.3%) | 297 (60.7%) |
| Air Pollution | 238 (33.0%) | 67 (28.9%) | 171 (35.0%) |
| Hearing Loss | 45 (6.2%) | 17 (7.3%) | 28 (5.7%) |
| Low Resilience | 301 (41.7%) | 61 (26.3%) | 240 (49.1%) |
| Low Literacy | 293 (40.6%) | 66 (28.4%) | 227 (46.4%) |

Continuous variables are reported as median (interquartile range), and categorical variables as n (%). Percentages were calculated using the full overall or diagnostic-group denominator, including participants with missing exposure data, and therefore do not represent assessed-only exposure prevalence. Cognitive and functional measures were not imputed (MoCA n=697; CDR-SB n=708; IDEA-IADL n=601). CU, cognitively unimpaired; MoCA, Montreal Cognitive Assessment; CDR-SB, Clinical Dementia Rating–Sum of Boxes (range 0-18); IDEA-IADL, Identification and Intervention for Dementia in Elderly Africans Instrumental Activities of Daily Living (range 0-33).

Dementia cases had higher recorded frequencies of depression (48.5% vs 32.3%), poverty (47.9% vs 22.0%), low resilience (49.1% vs 26.3%), and low literacy (46.4% vs 28.4%) than CU participants. Living alone, the operational measure of social isolation, was less common among dementia cases (4.3% vs 9.9%). These percentages use all participants in each diagnostic group as denominators, including those without a recorded value for a given exposure; they are not necessarily prevalences among participants who were actually assessed.

Missing values were especially common for obesity (576/721, 79.9%), PM₂.₅ exposure (430/721, 59.6%), and hypertension (284/721, 39.4%). The substantial missingness for obesity and hypertension reflects their introduction after the initial phase of data collection, whereas PM₂.₅ monitoring was not undertaken in all participants. Low literacy was missing for 125 participants (17.3%) and sleep-disorder history for 81 (11.2%). The complete distribution of observed and missing values is provided in Supplementary Table S1.

### Incremental model fit

Adding the complete 16-factor block improved Nagelkerke pseudo-R² from 0.1059 for the baseline model (with age and sex only) to 0.3682 for the full model (with age, sex, and 16 factors), a difference of ΔR²=0.2623 (bootstrap 95% CI 0.2414–0.3705; Table 4). The likelihood-ratio test statistic was χ²(27)=163.62 (p=1.66×10⁻²¹), demonstrating a significant improvement in model fit after including these 16 factors. Model parameter counts, including the intercept, were 1 for the intercept-only model, 3 for the baseline model (M0), and 30 for the full model (M1). The improvement of ΔR²=0.2623 measures additional in-sample model fit and cannot be interpreted as an attributable fraction or a change in population dementia prevalence.

### Adjusted associations

In the simultaneous model (Table 3; Figure 2), higher educational attainment (aOR 0.51, 95% CI 0.39–0.66) was associated with lower dementia odds. Poverty (aOR 2.34, 95% CI 1.52–3.60) and low resilience (aOR 1.84, 95% CI 1.18–2.86) were associated with higher dementia odds. Obesity was inversely associated with dementia (aOR 0.31, 95% CI 0.10–0.90); however, this association must be interpreted against nearly 80% missing obesity information and the potential for reverse causation and selection bias.

**Figure 2.**
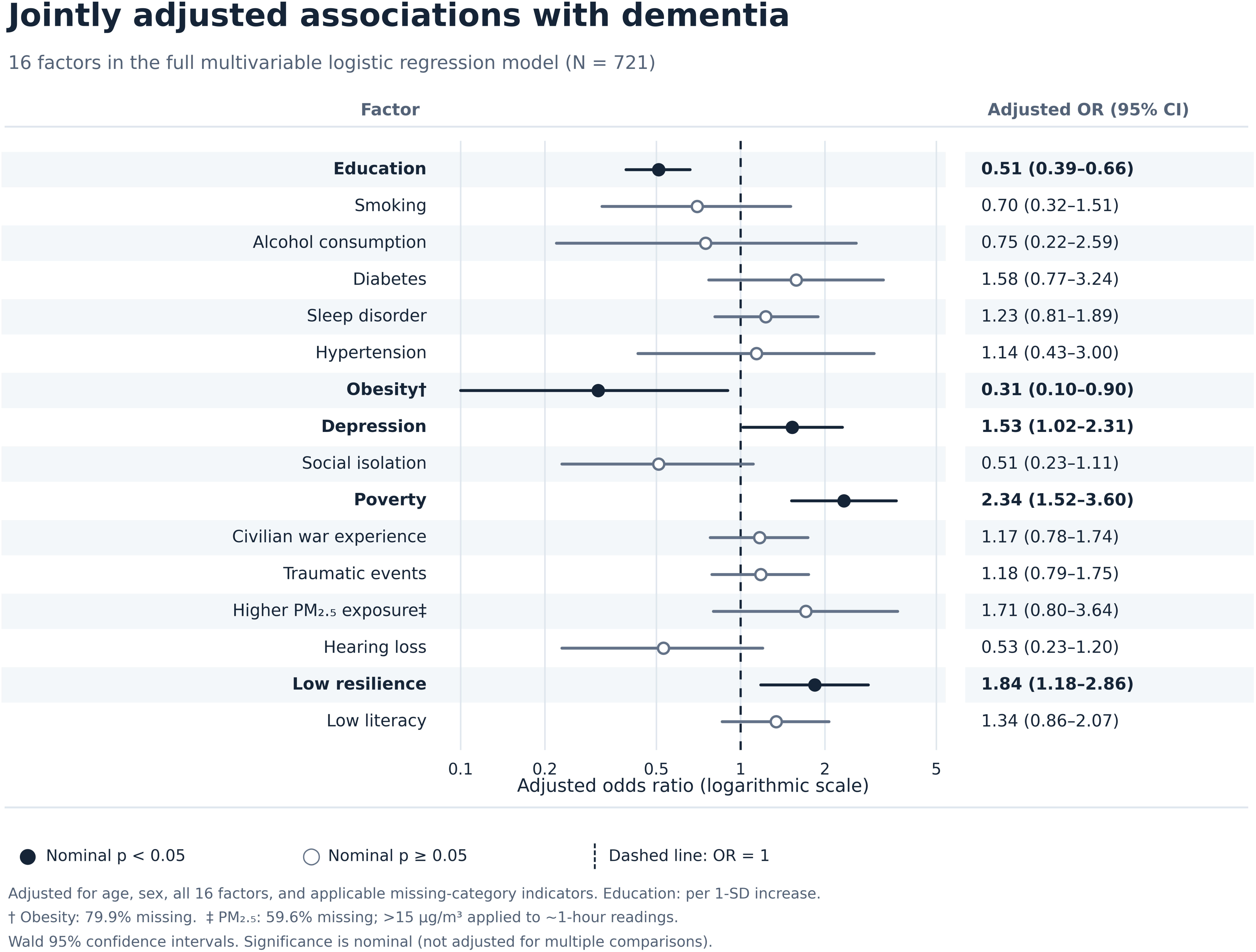
Adjusted odds ratios for 16 factors associated with dementia classification. Point estimates and horizontal lines represent adjusted odds ratios (aORs) and corresponding 95% Wald confidence intervals from the full multivariable logistic regression model (N=721). Estimates are displayed on a logarithmic scale, with the dashed vertical line indicating an odds ratio of 1. Filled circles indicate nominal statistical significance (p<0.05), whereas open circles indicate nonsignificant associations (p≥0.05). Estimates were adjusted for age, sex, all 16 factors, and relevant missing-category indicators. Education was modeled per standard-deviation increase. Obesity and PM₂.₅ measurements had substantial missingness (79.9% and 59.6%, respectively). Statistical significance was not adjusted for multiple comparisons. The PM₂.₅ cutoff of 15 µg/m³ was applied to approximately one-hour measurements.

**Table 3.** Adjusted odds ratios and corresponding 95% confidence intervals (CI) of the 16 factors with the dichotomous dementia outcome that were estimated by the full logistic regression model.

| **Modifiable Factors** | **Adjusted Odds Ratio (95% CI)** |
|---|---|
| Education | 0.51 (0.39, 0.66) |
| Smoking | 0.70 (0.32, 1.51) |
| Alcohol Consumption | 0.75 (0.22, 2.59) |
| Diabetes | 1.58 (0.77, 3.24) |
| Sleep Disorder | 1.23 (0.81, 1.89) |
| Hypertension | 1.14 (0.43, 3.00) |
| Obesity | 0.31 (0.10, 0.90) |
| Depression | 1.53 (1.02, 2.31) |
| Social Isolation | 0.51 (0.23, 1.11) |
| Poverty | 2.34 (1.52, 3.60) |
| War Experience | 1.17 (0.78, 1.74) |
| Traumatic Events | 1.18 (0.79, 1.75) |
| Air Pollution | 1.71 (0.80, 3.64) |
| Hearing Loss | 0.53 (0.23, 1.20) |
| Low Resilience | 1.84 (1.18, 2.86) |
| Low Literacy | 1.34 (0.86, 2.07) |

Estimates come from M1 (N=721), adjusted for age, sex, all 16 factors, and missing-category indicators. Education is per standard-deviation increase; binary factors compare “Yes” with “No.” Education, obesity, depression, poverty, and low resilience met nominal p<0.05 (uncorrected). Obesity data were 79.9% missing; its inverse association does not establish protection.

**Table 4.**
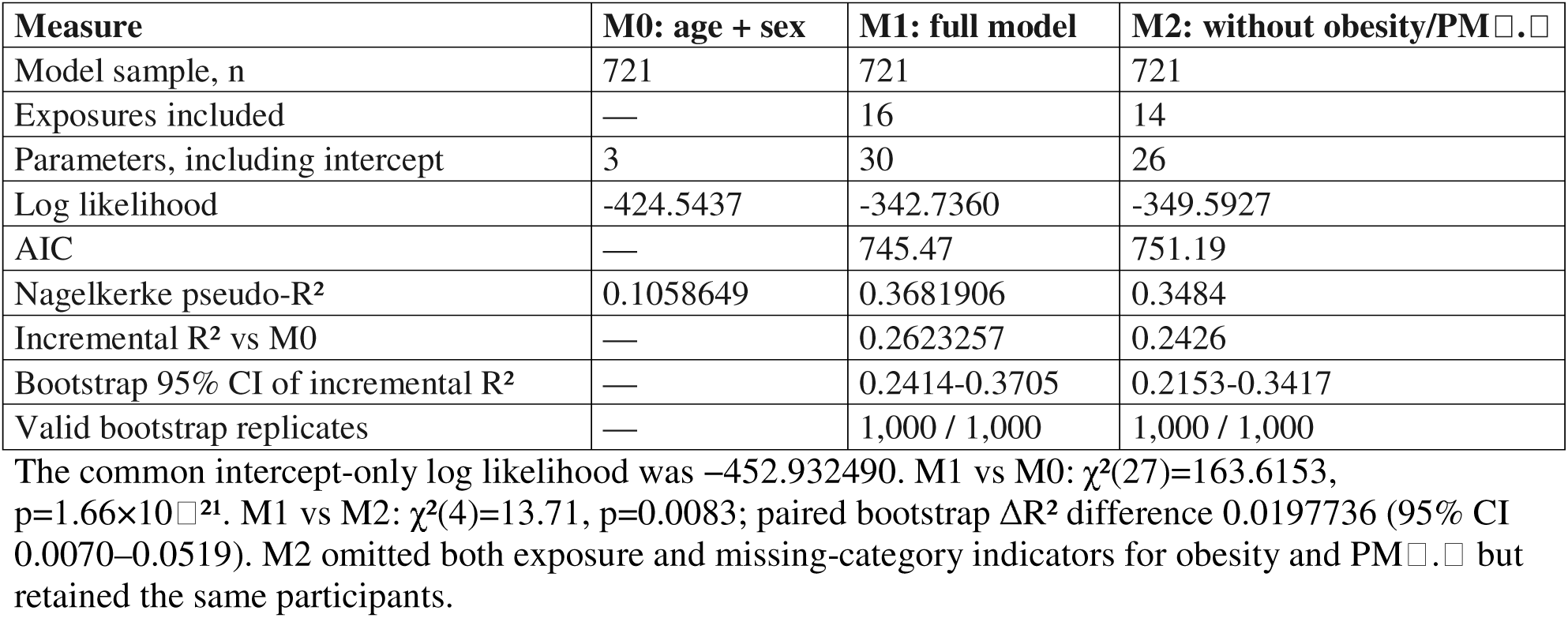
Nested logistic model fit and sensitivity analysis.

| Measure | M0: age + sex | M1: full model | M2: without obesity/PM[illegible].[illegible] |
|---|---|---|---|
| Model sample, n | 721 | 721 | 721 |
| Exposures included | — | 16 | 14 |
| Parameters, including intercept | 3 | 30 | 26 |
| Log likelihood | -424.5437 | -342.7360 | -349.5927 |
| AIC | — | 745.47 | 751.19 |
| Nagelkerke pseudo-R<sup>2</sup> | 0.1058649 | 0.3681906 | 0.3484 |
| Incremental R<sup>2</sup> vs M0 | — | 0.2623257 | 0.2426 |
| Bootstrap 95% CI of incremental R<sup>2</sup> | — | 0.2414-0.3705 | 0.2153-0.3417 |
| Valid bootstrap replicates | — | 1,000 / 1,000 | 1,000 / 1,000 |

The common intercept-only log likelihood was −452.932490. M1 vs M0: χ<sup>2</sup>(27)=163.6153, p=1.66×10<sup>[illegible]21</sup>. M1 vs M2: χ<sup>2</sup>(4)=13.71, p=0.0083; paired bootstrap ΔR<sup>2</sup> difference 0.0197736 (95% CI 0.0070–0.0519). M2 omitted both exposure and missing-category indicators for obesity and PM[illegible].[illegible] but retained the same participants.

Depression was associated with higher dementia odds (aOR 1.53, 95% CI 1.02–2.31), meeting the nominal significance threshold (p=0.0400). The confidence intervals for smoking, alcohol consumption, diabetes, sleep disorder, hypertension, social isolation, war experience, traumatic events, higher PM₂.₅, hearing loss, and low literacy all included 1.0. None of these results establishes a causal relationship with dementia. These adjusted odds ratios from the full logistic regression model reflect adjustment for age, sex, the other exposures, and relevant missing-category indicators.

### Missingness indicators and sensitivity analysis

Several missing-category indicators in the full model were associated with dementia status: obesity missingness (OR 0.36, 95% CI 0.19–0.68), sleep disorder missingness (OR 0.42, 95% CI 0.21–0.86), and low-literacy missingness (OR 0.52, 95% CI 0.30–0.91). These associations indicate that missingness may not be independent of dementia classification (Supplementary Table S2).

In the sensitivity model M2 excluding obesity, PM₂.₅, and their corresponding missing-category indicators, we retained 14 factors and 721 participants and calculated Nagelkerke pseudo-R²=0.3484, with ΔR²=0.2426 compared with M0 (bootstrap 95% CI 0.2153–0.3417). Comparing M1 with M2 using a likelihood-ratio test, we obtained χ²(4)=13.71 (p=0.0083); the paired bootstrap difference in ΔR² was 0.0198 (95% CI 0.0070–0.0519). These results indicate that the excluded exposure and missingness indicators contributed to model fit, although the difference was small and does not isolate the individual effects of obesity or PM₂.₅.

## DISCUSSION

In this community-derived cross-sectional sample of older adults in the DRC, the simultaneous inclusion of 16 social, psychosocial, behavioral, cardiometabolic, and environmental factors as covariates in the full multiple variable logistic regression model of dementia significantly improved the goodness of fit beyond the baseline model considering only age and sex. Particularly, higher poverty, depression, and lower resilience were associated with greater odds of dementia, whereas higher education was associated with lower odds of dementia. These mutually adjusted associations highlight contextual correlates of cognitive health with considered 16 modifiable factors in Central Africa, but do not quantify causal risks.

The association between dementia and multidimensional poverty merits particular attention. Poverty in the DRC may encompass reduced educational opportunity, limited access to preventive and clinical services, material insecurity, chronic stress, nutritional adversity, and environmental conditions. These pathways could influence cognitive reserve and health throughout the life course, although the present data do not establish when deprivation occurred relative to cognitive decline. The adapted poverty measure also includes educational and health-related dimensions, meaning that its association cannot be cleanly separated from education, functioning, or other health indicators in the same model. Its adjusted odds ratio should therefore be interpreted as a marker of overlapping disadvantage, not the isolated effect of a single manipulable exposure.

The inverse association between years of education and dementia is consistent with literature emphasizing educational access and cognitive reserve in dementia epidemiology.^2,5,6^ Education may represent early-life social opportunity in addition to a putative cognitive reserve mechanism. Importantly, education was modeled continuously and standardized in the primary analysis, whereas a dichotomous “low education” measure featured in the original manuscript. Comparisons to previous low-education population attributable fractions are consequently not direct. Associations may also be influenced by the use of cohort-based age-, sex-, and education-adjusted cognitive assessment norms in diagnostic classification, which underscores the need for validation in representative samples.

Low resilience was associated with dementia status even after accounting for poverty, depression, trauma, and other factors. This observation raises questions about the interplay of psychosocial resources, late-life stress exposure, depressive symptoms, and cognitive or functional impairment. However, resilience was measured at the same baseline assessment as dementia and could itself be affected by cognitive decline, functional dependence, or reporting patterns. It should not be presented as evidence that resilience training would prevent dementia. Depression was also associated with higher dementia odds in the adjusted model, meeting the nominal significance threshold (aOR 1.53, 95% CI 1.02–2.31; p=0.0400). However, the cross-sectional design limits conclusions about the temporal relationship between depressive symptoms and dementia.

The inverse association between obesity and dementia warrants particularly cautious interpretation. Reverse causation, including prodromal weight loss, frailty, survivor bias, and age-related changes in body composition may complicate the relationship between late-life obesity and cognitive impairment.^17,18^ Importantly, obesity data were unavailable for approximately 80% of participants because obesity assessments were incorporated after the initial phase of data collection. This substantial missingness may introduce selection bias, particularly if participants enrolled during different study phases differed systematically in demographic or clinical characteristics. Similarly, hypertension assessments were introduced later in the study, and PM₂.₅ exposure was not measured in all participants. Although missing-data categories were included in the regression models, this approach does not eliminate potential bias arising from systematically unavailable measurements. Accordingly, the inverse association observed for obesity should not be interpreted as evidence of a protective effect.

Some anticipated factors, including diabetes, hypertension, smoking, hearing loss, war experience, and trauma exposure, did not show statistically distinguishable adjusted contrasts in the present model. Null or imprecise findings do not imply these factors are unimportant for dementia prevention. The combined analysis included correlated exposures and multiple missingness indicators; limited numbers in some observed exposure categories, outcome-enriched sampling, measurement error, and adjustment for related socioeconomic factors may all alter estimates. The self-reported war and trauma items did not comprehensively measure timing, duration, or severity of exposure. PM₂.₅ was measured using short-term indoor monitoring, which may inadequately approximate sustained exposure.^19^ The cutoff of 15 µg/m³ was applied to approximately one-hour measurements and should not be interpreted as evidence of exposure exceeding a 24-hour average guideline.

The present findings should be distinguished from population attributable fraction estimates reported in previous dementia prevention research. Although the evaluated factors collectively improved the goodness of fit of the logistic regression model of dementia, Nagelkerke pseudo-R² does not represent the proportion of dementia cases caused by these exposures or potentially preventable through their modification. Furthermore, because this study used cross-sectional data from a nonrepresentative, dementia-enriched sample, the observed associations cannot establish causality or be used to extrapolate preventable dementia burden to the broader DRC population. Direct comparisons with population-level prevention estimates, including those reported by the Lancet Commission,^2^ are therefore not appropriate. This distinction is particularly important because the sample is enriched for dementia and is not population-representative.

The public health relevance of the results lies in highlighting potential priorities for prospective investigation, not establishing intervention effect sizes. Poverty reduction, equitable access to education and health care, and psychosocial support may be worthy of consideration alongside established vascular risk strategies.^2,9,10^ Their independent and potentially mediated relationships with dementia warrant study using longitudinal trajectories, validated measurement, and analyses designed to distinguish causal effects from shared causes and reverse causation. Future work should evaluate clinically classified Alzheimer’s disease and other dementias separately when adequately powered.

Strengths include a locally implemented, community-engaged dementia cohort, culturally adapted cognitive and functional assessments, multidisciplinary clinical classification, broad coverage of conventional and context-specific factors, and transparent reporting of model fit and missing data.^11^ The use of the same participant sample for nested-model comparisons avoids variation in sample composition across M0 and M1, and the sensitivity model explicitly probes heavily missing exposure domains.

Limitations include cross-sectional measurement, which precludes establishing exposure timing or causality, and community/clinical recruitment, which may produce selection bias. The sample excludes MCI and combines clinically classified suspected Alzheimer’s disease with other dementias; Alzheimer’s disease was not biomarker-confirmed in this analysis. The published cohort description used age ≥65 years for recruitment, whereas the analytic extract was evaluated using an age-60 cutoff for known ages, and two missing ages were mean-imputed; the source of this eligibility discrepancy warrants clarification. Obesity and hypertension were added after the initial study phase, and PM₂.₅ measurements were not obtained from all participants, creating substantial and potentially informative missingness. Missing-category indicators and mean imputation do not eliminate missing-data bias, and fixed bootstrap preprocessing does not reflect uncertainty in imputed values. Correlated exposures, overlapping poverty-index dimensions, multiple testing, and lack of external validation further constrain interpretation. The model does not quantify the population burden attributable to any exposure.

### Conclusions

In this community-derived DRC cohort, poverty, low resilience, depression, and education showed mutually adjusted associations with dementia status, and adding the full factor block increased in-sample logistic model fit beyond age and sex. The results underscore the potential importance of structural and contextual exposures in future dementia research; they do not establish preventability or intervention effect sizes. Prospective studies with complete and standardized exposure data, transparent sample ascertainment, and more representative recruitment are needed.

## Acknowledgments

The authors thank the participants, their families and caregivers, community partners, and the clinical and research staff involved in the EVCD-RDC study. We also acknowledge the Centre Médical de Kinshasa for its support of study procedures and data collection.

## Author Contributions

Jean Ikanga: Conceptualization, Methodology, Investigation, Funding acquisition, Project administration, Supervision, Writing – original draft, Writing – review & editing.

Saranya Sundaram Patel: Writing – original draft, Writing – review & editing.

Peng Zhang: Formal analysis.

Jingjing Yang: Formal analysis.

Caterina Obenauf: Project administration, Writing – review & editing.

Guy Gikelekele: Investigation.

Emmanuel Epenge: Investigation.

Japhet Magolu Potshi: Investigation.

Topina Tomadia: Investigation.

Joseph Phuati Tsangu: Investigation.

Samuel Mampunza: Investigation.

Lelo Mananga: Investigation.

Justine Bukabau: Investigation.

Thomas Karikari: Writing – review & editing.

Alden L. Gross: Writing – review & editing.

Alvaro Alonso: Writing – review & editing.

## Statements and Declarations

### Ethical Considerations

The Étude du Vieillissement Cognitif et de Démence en République Démocratique du Congo (EVCD-RDC) study protocol was approved by the Research Ethics Committee of the School of Public Health at the University of Kinshasa (original protocol ESP/CE/054c/2024; approval renewed July 18, 2026, through July 17, 2027) and the Emory University Institutional Review Board (IRB ID MOD006-IRB00113463). Study procedures were conducted in accordance with applicable ethical standards and the Declaration of Helsinki.

### Consent to Participate

Written informed consent was obtained before participation in the parent EVCD-RDC study. As described in the study protocol, consent could be provided independently or, when applicable, through a caregiver or appropriately authorized representative in accordance with institutional ethics requirements.

### Consent for Publication

Not applicable.

### Data Availability

Deidentified participant-level data underlying the findings of this study, together with an accompanying data dictionary where available, may be made available to qualified researchers upon reasonable request to the corresponding author. Data access is subject to approval by the EVCD-RDC study investigators and relevant ethics committees, applicable participant-consent and institutional restrictions, and completion of a data-use agreement.

## References

1. Nichols E, Steinmetz JD, Vollset SE, et al. Estimation of the global prevalence of dementia in 2019 and forecasted prevalence in 2050: an analysis for the Global Burden of Disease Study 2019. Lancet Public Health 2022; 7: e105–e125. doi:10.1016/S2468-2667(21)00249-8.

2. Livingston G, Huntley J, Liu KY, et al. Dementia prevention, intervention, and care: 2024 report of the Lancet standing Commission. Lancet 2024; 404: 572–628. doi:10.1016/S0140-6736(24)01296-0.

3. Mukadam N, Sommerlad A, Huntley J, et al. Population attributable fractions for risk factors for dementia in low-income and middle-income countries: an analysis using cross-sectional survey data. Lancet Glob Health 2019; 7: e596–e603. doi:10.1016/S2214-109X(19)30074-9.

4. Stephan BCM, Cochrane L, Kafadar AH, et al. Population attributable fractions of modifiable risk factors for dementia: a systematic review and meta-analysis. Lancet Healthy Longev 2024; 5: e406–e421. doi:10.1016/S2666-7568(24)00061-8.

5. Oliveira D, Otuyama LJ, Mabunda D, et al. Reducing the number of people with dementia through primary prevention in Mozambique, Brazil, and Portugal: an analysis of population-based data. J Alzheimers Dis 2019; 70(Suppl 1): S283–S291. doi:10.3233/JAD-180636.

6. Ojagbemi A, Okekunle AP and Babatunde O. Dominant and modifiable risk factors for dementia in sub-Saharan Africa: a systematic review and meta-analysis. Front Neurol 2021; 12: 627761. doi:10.3389/fneur.2021.627761.

7. Sixsmith J, Sixsmith A, Callender M, et al. Wartime experiences and their implications for the everyday lives of older people. Ageing Soc 2014; 34: 1457–1481. doi:10.1017/S0144686X13000214.

8. Burnes DPR and Burnette D. Broadening the etiological discourse on Alzheimer’s disease to include trauma and posttraumatic stress disorder as psychosocial risk factors. J Aging Stud 2013; 27: 218–224. doi:10.1016/j.jaging.2013.03.002.

9. Mayston R, Frissa S, Tekola B, et al. Explanatory models of depression in sub-Saharan Africa: synthesis of qualitative evidence. Soc Sci Med 2020; 246: 112760. doi:10.1016/j.socscimed.2019.112760.

10. Angendu KB, Akilimali PZ, Tran NT, et al. Dynamic progression of hypertension and diabetes in the Democratic Republic of Congo from 2019 to 2023. J Clin Med 2024; 13(18): 5488. doi:10.3390/jcm13185488.

11. Ikanga J, Obenauf C, Patel SS, et al. Cohort profile: a community-based prospective cohort study of Alzheimer’s disease and related dementias in the Democratic Republic of Congo. Alzheimers Dement Behav Socioecon Aging 2026; 2(2): e70085. doi:10.1002/bsa3.70085.

12. Ikanga J, Reyes A, Kaba D, et al. Prevalence of suspected dementia in a sample of adults living in Kinshasa-Democratic Republic of the Congo. Alzheimers Dement 2023; 19(9): 3783–3793. doi:10.1002/alz.13003.

13. Paddick SM, Gray WK, Ogunjimi L, et al. Validation of the Identification and Intervention for Dementia in Elderly Africans (IDEA) cognitive screen in Nigeria and Tanzania. BMC Geriatr 2015; 15: 53. doi:10.1186/s12877-015-0040-1.

14. Manly JJ, Jones RN, Langa KM, et al. Estimating the prevalence of dementia and mild cognitive impairment in the US: the 2016 Health and Retirement Study Harmonized Cognitive Assessment Protocol Project. JAMA Neurol 2022; 79(12): 1242–1249. doi:10.1001/jamaneurol.2022.3543.

15. Besser L, Kukull W, Knopman DS, et al. Version 3 of the National Alzheimer’s Coordinating Center’s Uniform Data Set. Alzheimer Dis Assoc Disord 2018; 32(4): 351–358. doi:10.1097/WAD.0000000000000279.

16. Nagelkerke NJD. A note on a general definition of the coefficient of determination. Biometrika 1991; 78(3): 691–692. doi:10.1093/biomet/78.3.691.

17. Natale G, Zhang Y, Hanes DW, et al. Obesity in late life as a protective factor against dementia and dementia-related mortality. Am J Alzheimers Dis Other Demen 2023; 38: 15333175221111658. doi:10.1177/15333175221111658.

18. Wu Y, Li D and Vermund SH. Advantages and limitations of the body mass index (BMI) to assess adult obesity. Int J Environ Res Public Health 2024; 21(6): 757. doi:10.3390/ijerph21060757.

19. Katoto PDMC, Byamungu L, Brand AS, et al. Ambient air pollution and health in sub-Saharan Africa: current evidence, perspectives and a call to action. Environ Res 2019; 173: 174–188. doi:10.1016/j.envres.2019.03.029.

