## Supplementary Table for "Modifiable and Context-Specific Factors Associated with Dementia in the Democratic Republic of the Congo: A Community-Derived Cross-Sectional Study"

### Supplementary Materials

#### Supplementary Table S1. Observed and missing counts in final analytic sample

| Variable | CU observed / missing | Dementia observed / missing | Total missing |
| --- | --- | --- | --- |
| Age | 232 / 0 | 487 / 2 | 2 (0.3%) |
| Sex | 232 / 0 | 489 / 0 | 0 (0.0%) |
| Education | 232 / 0 | 488 / 1 | 1 (0.1%) |
| Smoking | 224 / 8 | 450 / 39 | 47 (6.5%) |
| Alcohol consumption | 230 / 2 | 477 / 12 | 14 (1.9%) |
| Diabetes | 207 / 25 | 466 / 23 | 48 (6.7%) |
| Hypertension | 139 / 93 | 298 / 191 | 284 (39.4%) |
| Obesity | 28 / 204 | 117 / 372 | 576 (79.9%) |
| Depression | 232 / 0 | 489 / 0 | 0 (0.0%) |
| Social isolation | 232 / 0 | 489 / 0 | 0 (0.0%) |
| Sleep disorder | 190 / 42 | 450 / 39 | 81 (11.2%) |
| High PM2.5 | 87 / 145 | 204 / 285 | 430 (59.6%) |
| Hearing loss | 230 / 2 | 478 / 11 | 13 (1.8%) |
| War experience | 232 / 0 | 489 / 0 | 0 (0.0%) |
| Traumatic experience | 232 / 0 | 489 / 0 | 0 (0.0%) |
| Low resilience | 214 / 18 | 450 / 39 | 57 (7.9%) |
| Low literacy | 184 / 48 | 412 / 77 | 125 (17.3%) |
| Poverty | 217 / 15 | 449 / 40 | 55 (7.6%) |

Final analytic n=721 (232 CU; 489 dementia). See revised analyses for 13 excluded dementia cases with incomplete data for depression, social isolation, war exposure, or traumatic events.

#### Supplementary Table S2. Associations of missing-category indicators with dementia

| Variable | Missing vs No OR (95% CI) | p-value |
| --- | --- | --- |
| Smoking | 1.56 (0.58-4.26) | 0.3807 |
| Alcohol Consumption | 4.73 (0.68-32.86) | 0.1161 |
| Diabetes | 0.46 (0.17-1.21) | 0.1163 |
| Hypertension | 0.87 (0.32-2.32) | 0.7744 |
| Obesity | 0.36 (0.19-0.68) | 0.0014 |
| Sleep Disorder | 0.42 (0.21-0.86) | 0.0172 |
| Air Pollution | 1.61 (0.78-3.33) | 0.2007 |
| Hearing Loss | 2.92 (0.53-15.97) | 0.2172 |
| Low Resilience | 1.25 (0.58-2.71) | 0.5651 |
| Low Literacy | 0.52 (0.30-0.91) | 0.0216 |
| Poverty | 2.11 (0.98-4.56) | 0.0571 |

Estimates adjusted for age, sex, and all full-model exposure and missing-category terms (M1). Missingness categories are not substantive exposure effects.

### Supplementary methodological details

The analyst-derived results use the 721-person final sample (232 CU and 489 dementia) and the same modeling and missingness-indicator strategy for the full and sensitivity models. The cohort’s published recruitment criterion (≥65 years) differs from the analytic extract’s known-age exclusion rule (<60 years); the latter was confirmed by the principal investigator.

Continuous variables were imputed using cohort means before standardization; age and education were standardized. Binary variables were represented by “Yes” and, where necessary, “Missing” indicators, with “No” as reference. Imputation and standardization were held fixed in bootstrap replicates. The bootstrap was used for within-sample changes in Nagelkerke pseudo-R², not external validation.

Sensitivity model M2 removed obesity and PM₂.₅ exposure and missing-category terms, while retaining the same 721 participants. The observed full-versus-sensitivity ΔR² difference was 0.0197736, with 95% paired bootstrap interval 0.0070–0.0519 and 1,000 valid paired samples.
